# A rapid One-Pot RNA-isolation method for simplified clinical detection of SARS-COV-2 infection in India

**DOI:** 10.1101/2022.05.26.22275661

**Authors:** Sonia Jain, Arghya Bhowmick, Anil Kumar Pandey

## Abstract

**Background:** With the rapid increase in COVID-19 cases and the discovery of new viral variants within India over multiple waves, the expensive reagents and time-consuming sample pretreatment required for qPCR analysis have led to slower detection of the disease. The vast Indian population demands an inexpensive and competent sample preparation strategy for rapid detection of the disease facilitating early and efficient containment of the disease.

**Methods:** In this study, we have surveyed the spread of COVID-19 infection over Faridabad, Haryana, India for 6 months. We also devised a simple single-step method for total RNA extraction using a single tube and compared its efficacy with the commercially available kits.

**Findings:** Our findings suggest that determining Ct values for samples subjected to the One Pot (OP) RNA extraction method was as efficient as the commercially available kits but delivers a subtle advantage in a way, by minimizing the cost, labor, and sample preparation time.

**Conclusion:** This novel crude RNA extraction method is stable and capable of operating in developing countries like India for low resource settings, without the use of expensive reagents and instruments. Additionally, this method can be further adapted to pooling samples strategies owing to its high sensitivity.

## Introduction

Coronaviruses (CoVs) existed throughout human history, and are known to cause mild respiratory illnesses in humans (Ye *et al*., 2020). The co-evolution with the human immune system bestows the CoVs capability of evading the protective responses and causing severe infection (Schultze and Aschenbrenner, 2021). In December 2019, a new member of the CoV family emerged in China; later termed as Severe Acute Respiratory Syndrome (SARS) associated CoV-2. SARS-CoV-2 causes a severe form of respiratory illness called COVID-19 (World Health Organization. Office of Library and Health Literature Services., 1988). In contrast to SARS or Middle East Respiratory Syndrome (MERS), the SARS-CoV-2 has shown unpredicted spread over the world where 4 million individuals (approximately) contracted the infection with more than 281736 fatalities (WHO Situation Report, May 2020) (World Health Organization, 2020). The current COVID 19 pandemic due to widespread human-to-human transmission is now rising at an alarming rate (World Health Organization. Office of Library and Health Literature Services., 1988). The variable immune response in the host has displayed a spectrum of clinical patterns ranging from mild to severe cases (García, 2020). Besides symptomatic cases, the increase in the number of cases may be due to the contact with the asymptomatic individual leading to rapid spread in the community (Oran and Topol, 2020). Therefore, to control this spreading of the infections from one community to another and to provide accurate treatment to patients, correct diagnosis of the pathogen is the first-line of requirement. Although many different diagnostic approaches have been introduced, the development of a rapid, inexpensive, and highly sensitive approach for clinical diagnosis remains a challenge (Kellner *et al*., 2019; Paz *et al*., 2020; Azhar *et al*., 2021). Its effective implementation would offer immense possibilities for rapid and widespread testing that has so far proven to be necessary for measuring the progression of the disease in multiple countries. The quantitative Reverse Transcription-Polymerase Chain Reaction (qRT-PCR) has been widely used by several laboratories as a gold standard test for routine diagnosis of COVID-19 infection, developed by the USA-CDC (Carter *et al*., 2020). However, inadequate access to reagents and equipment due to high cost and the time-consuming sample pre-treatment has slowed the disease detection and impeded efforts to prevent viral spread (Vandenberg *et al*., 2021). With clinical samples, fast, inexpensive and sensitive RNA isolation methods may help in point-of-care (POC) pathogen detection. The objective of this study was to develop a simple, rapid and inexpensive one-tube testing method to diagnose SARS-CoV-2 which can be performed in low-resource settings.

## Materials and Methods

### Study Population

The study was carried out in ESIC Medical college and hospital, Faridabad, Haryana, India **(Figure 1)**. In this study, we surveyed the COVID-19 infections spread over Haryana for a period of 6 months (May 2020 to October 2020). This study was approved by the Indian government and the institutional review boards at participating institutions. Patients with severe acute respiratory illness (SARI) or influenza like illness (ILI) i.e., clinically symptomatic population are recruited as per the guidelines of the Institutional Ethics Committee. The demographic characteristics like age, gender, travel history like migrant laborers, etc., place of residence, source of income, and co-morbidities if any was determined by taking a detailed history of the patient. The study excluded those individuals who refused to give written consent. Also, hospitalized patient or children with symptoms not related to COVID-19 infection was excluded from this study.

**Figure 1:**
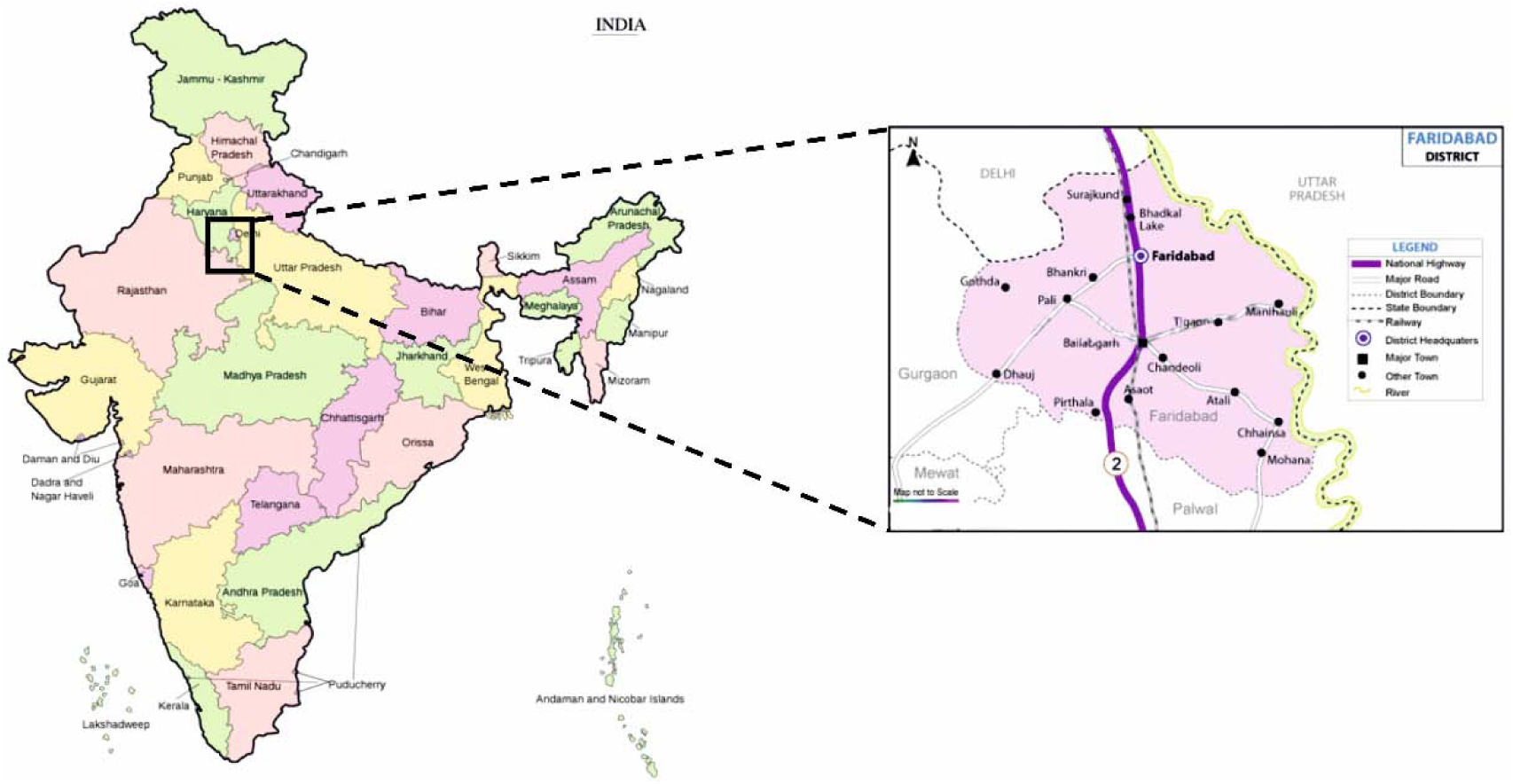
Geographical map of India showing the exact location of Faridabad district in the state of Haryana, where this study was carried out.

### Sample collection and transportation

Before taking samples, informed consent was obtained from the patients or parents/guardians of the children. The samples were collected according to ICMR 2020 COVID-19 test protocol. Nasopharyngeal (NP) and oropharyngeal (OP) swab samples were collected from the consecutive population using sterile nylon flocked swabs and were immediately placed in viral transport medium (VTM), labeled and transported on the ice at the earliest to the laboratory for the detailed investigations.

### RT-PCR for different genes of SARS-Cov-2

For the qRT-PCR analysis, the kits approved by ICMR (Indian Council of Medical Research) were employed for preparing samples as per the manufacturer’s instructions. The amplification of SARS-COV-2 genes was carried out by targeting the, (*N*) Nucleocapsid gene, (*E*) Envelope gene, (*RdRp*) RNA-dependent RNA polymerase gene (Udugama *et al*., 2020; Cho *et al*., 2020). The samples with a Cycle threshold (Ct) value ≤ 40 were interpreted as positive, while Ct value >40 or N.A. were interpreted as negative. The samples which were positive for *N* gene, *E* gene and *RdRp* gene after qRT-PCR analysis were considered as COVID-19 positive. Each time the qRT-PCR results were validated by positive amplification of the Internal Control (IC) gene provided in the kit.

### Study Design

It is a retrospective study where out of a total of 58252 cases, only 500 cases were selected; 250 NP/OP aliquots from a previously tested COVID-19 positive group of individuals and 250 NP/OP aliquots from previously tested COVID-19 negative group of individuals were taken **(Figure 2)**. The viral RNA was extracted from all these NP/OP swabs by employing our newly devised One-Pot (OP) RNA isolation method. The positive and negative results for SARS-COV-2 virus genes were determined by the Ct value obtained from qRT-PCR tests, using kits validated by ICMR.

**Figure 2:**
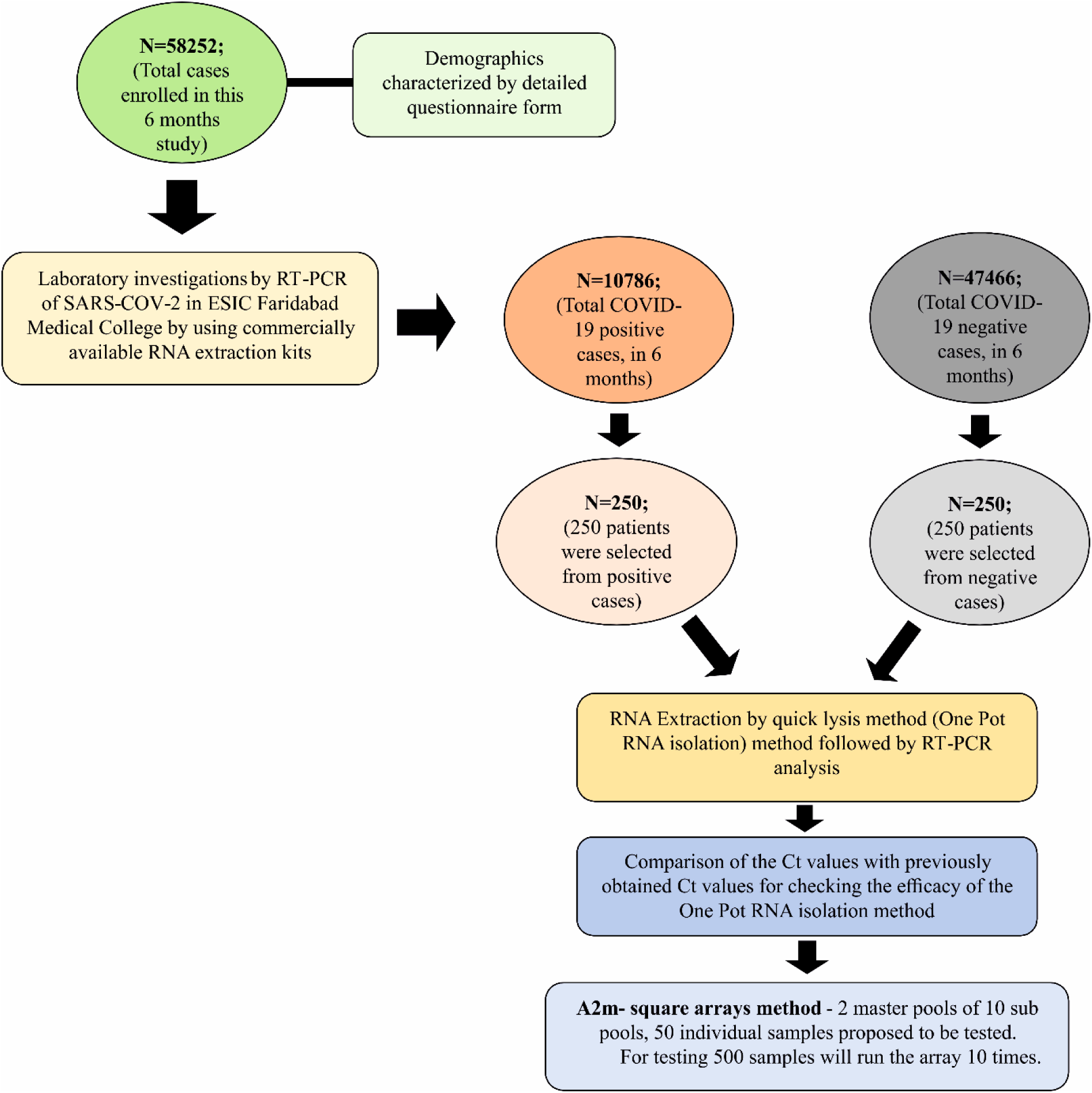
Flow diagram depicting the study design employed in carrying out this work.

### RNA Extraction by quick lysis method

The previous Acid-guanidinium-phenol-chloroform (AGPC) method of RNA extraction by Chomczynki and Sacchi was further modified for extracting viral RNA from the patient’s nasopharyngeal swab samples (CHOMZYNSKI, 1987). We devised a single tube quick-lysis method for a fast and efficient yield of crude viral RNA. This One-Pot (OP) RNA extraction method includes the use of the following reagents: a) Quick Lysis solution (4M guanidinium thiocyanate, 0.5% (wt/vol) *N-*lauryl sarcosine, b) Chloroform, c) Glycogen solution (3.5mg/ml stock) and d) 100% cold ethanol.

Several microcentrifuge tubes (2ml) containing 500 μl Quick-Lysis solution and 300 μl chloroform were kept as ready-to-use stocks. While testing, 200 μl of NP/OP swab from VTM (Viral transport media) was directly added to the 800 μl of ready-to-use stock in the microcentrifuge tube followed by the addition of 5 μl of glycogen solution. The mixture was then vortexed and incubated at 65ºC for 15 mins. Further, it was centrifuged at 14000 rpm for 5 mins allowing phase separation **(Figure 3)**. The upper yellow layer was discarded and 1 ml of ice-cold ethanol solution was added to the bottom layer followed by again centrifugation at 14000 rpm for 5 mins. Finally, the supernatant was discarded followed by air drying of the tubes for 10 mins (to remove residual ethanol). The RNA pellet was then further resuspended in 20 μl of DEPC-treated water, quantified and subsequently used as a template in qRT-PCR analysis for detecting viral load in NP/OP swabs of affected patients. The Cycle threshold (Ct) values obtained after extracting RNA using this method were compared to the Ct values attained using a commercial RNA isolation kit. We performed non-parametric Mann Whitney tests to determine whether differences between the Ct values of the sample prepared using a commercial RNA extraction kit and the new One-pot RNA isolation method were significant for the three genes tested. Significance level was based on 2-tailed α (significance level) = 0.05.

**Figure 3:**
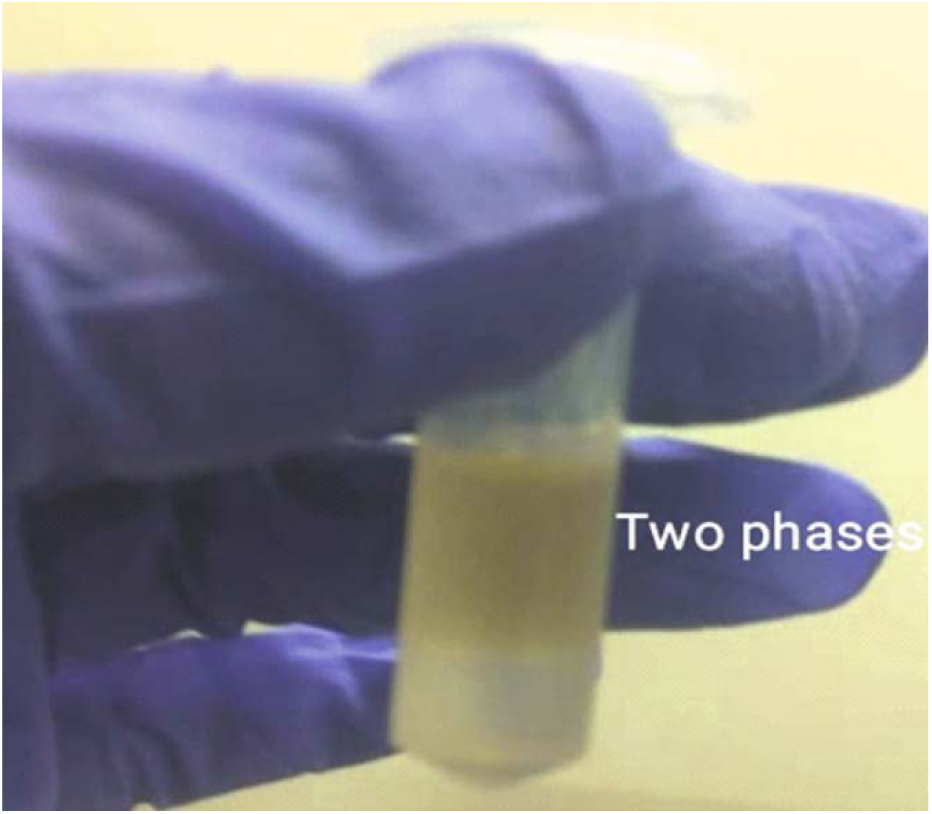
A microfuge tube showing two-phase separation after addition of Quick lysis solution, chloroform, and glycogen to VTM sample containing the NP/OP swab followed by brief heating at 65 □ and subsequent centrifugation at high speed.

### Pooling testing strategies

The Ministry of Health and Family Welfare, Govt. of India has made provision for a one-time pool sample of 25 to be tested for Covid-19 district-level surveillance. Therefore, the modified A2m-square arrays method with two different master pools was applied in this study which generally provides better efficiency and positive predictive value (PPV) (Taylor *et al*., 2010). This pooling strategy is a two-dimensional array (“A2m”) with master pool testing. The first test was carried out for the master pool; if the master pool tests positive, then pools consisting of all samples in each row and column of the array are tested at the same time. Retesting is done for all the samples at points of intersection between positive rows and positive columns. In the situation of a positive row but no positive columns, all samples in the positive row are tested. As testing row and column pools occur in a single step of the testing process, thus A2m is a three-stage pooling algorithm. Only A2m algorithms with the similar number of rows and columns will be taken into account. In testing this algorithm, the number of samples included in each pool depends on the expected prevalence rate of the disease under study and the characteristics of the diagnostic test. Further, it also depends on dilution effects in the master pool, the length of the sensitivity depends on both the sensitivity/ lower limit of detection of the assay and viral load in acute infection (Westreich *et al*., 2008). The optimal pool size required for testing was calculated with an available online calculator (http://www.bios.unc.edu/_mhudgens/optimal.pooling.b.htm). Therefore, according to the pooling size strategy mentioned above, the master pools size was 25. Two master pools of 10 sub-pools and 50 individual samples were tested in this algorithm. So, for testing 500 samples we have to run the array 10 times. For quality control, 5% of negative samples were randomly tested by standard qRT-PCR to check the false-negative rate.

## Results

### COVID-19 prevalence over Faridabad, Haryana, India

For the period of 6 months, a total of n=58252 samples were collected from different individuals who came to visit ESIC Medical college and hospital, Faridabad, Haryana for routine clinical checkups and testing. The RT-PCR testing of samples collected from their NP/OP swabs indicated 10786 individuals were positive for COVID-19 while the rest of 47466 were tested negative for COVID-19 indicating a prevalence rate of 18.52%. Out of the total n=58252 samples: 35184 samples were from males; 22805 samples were collected from females while 262 samples were from neonates. A total of 7086 males were found to be positive for COVID-19 while 3700 females were positive for COVID-19 **(Table 1)**. However, all the neonates tested negative for COVID-19. The percentage of occurrence of COVID-19 was calculated to be 20.1% and 16.2% in the case of males and females respectively for the period of 6 months of study. The infected percentage obtained **(Figure 4a)** for the different age groups are as follows: 0-17 years – 9.7%; 18-35 years-16.2%; 36-53 years-23.8%; 54-71 years-25.4% and 72-89 years-28.8% **(Table 1)**. Thus, our data indicate that SARS-CoV-2 is highly affecting individuals belonging to higher age groups. Among those who were positive for SARS-COV-2, the most predominant comorbid conditions were diabetes (37.2%), hypertension (30%), Respiratory disorder (24%), renal disorder (14.7 %) and obesity (11.2%) **(Table 1)**. Amongst the COVID-19 affected patients, most of the patients had Ct values within 25.01 to 35.00 for the *N* gene **(Figure 4b)** while, Ct values within 20.01 to 35.00 were observed for both the *RdRp* and *E* gene. Thus, from our data, we can speculate that the relative abundance of the viral *RdRp* and *E* gene is much higher compared to the *N* gene in the affected patient samples.

**Table 1:**
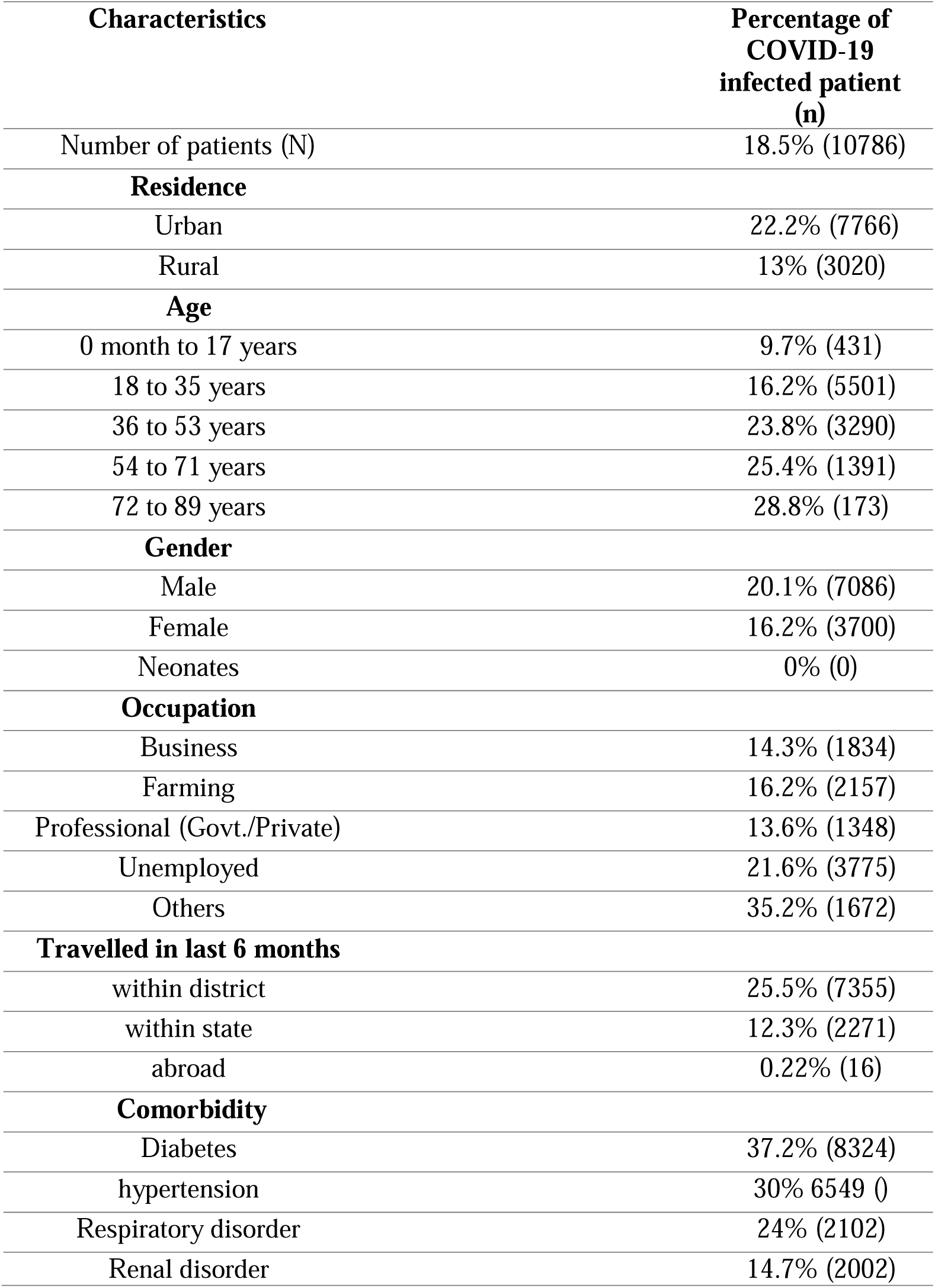
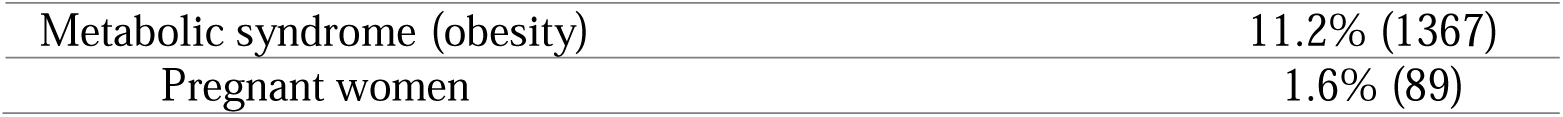
Table showing the percentage of COVID-19 infected patients and the total number of COVID-19 infected patients (n) in each of the different groups. The percentage of COVID-19 infected patients is calculated by dividing the total COVID-19 positive patients in the group by total patients registered in that corresponding group followed by multiplying by 100.

**Figure 4:**
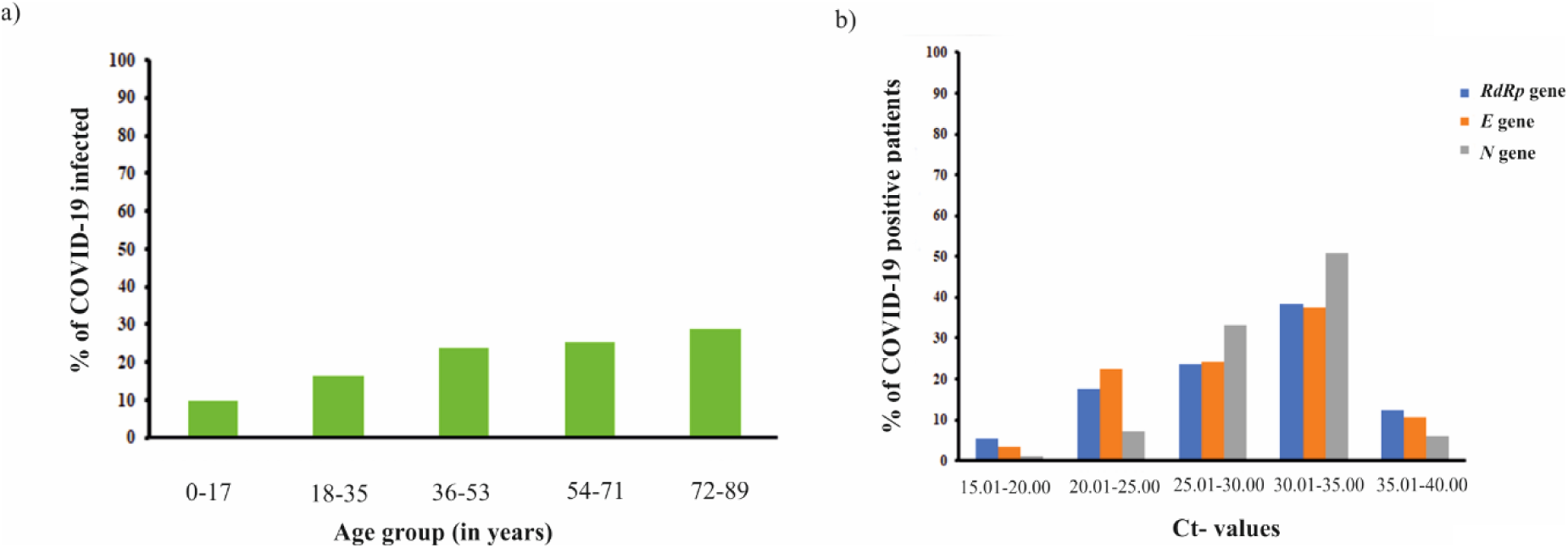
**a)** Bar graph representation showing the percentage of COVID-19 infected patients from different age groups. The percentage of COVID-19 infected patients raised with the increase in the age group of the individuals **b)** Graph showing correlation of Ct-values of different viral genes (*RdRp* gene, *E* gene and *N* gene) with the percentage of COVID-19 positive patients.

### One-pot (OP) RNA isolation method was effective in extracting viral RNA from NP/OP swabs

This One-pot (OP) RNA isolation method employed simple denaturing and a precipitation solution, with a centrifugation method to extract the crude RNA which can be directly used for RT-PCR testing. The Quick lysis solution was able to lyse the SARS-COV-2 Virus coating within a very short time. Also, heating at a higher temperature reduces the risk of handling positive samples without compromising the integrity of the viral RNA. Similar to other commercially available RNA isolation kits, this new RNA isolation method shows the Ct-value and Internal Control (IC) perfectly. The Ct values obtained after RT-PCR analysis using this new method of RNA isolation were almost similar to those obtained after RT-PCR analysis utilizing the commercial method of RNA isolation **(Figure 5)**. Previously tested samples negative for COVID-19 were found to be negative (data not shown) and COVID-19 positive samples were positive with this new RNA isolation method **(Figure 5)**. We have also performed the pooling strategy where samples were pooled and then tested. The positive result obtained from the highly diluted pooled samples marked the uncompromised sensitivity of this newly devised RNA-isolation method.

**Figure 5:**
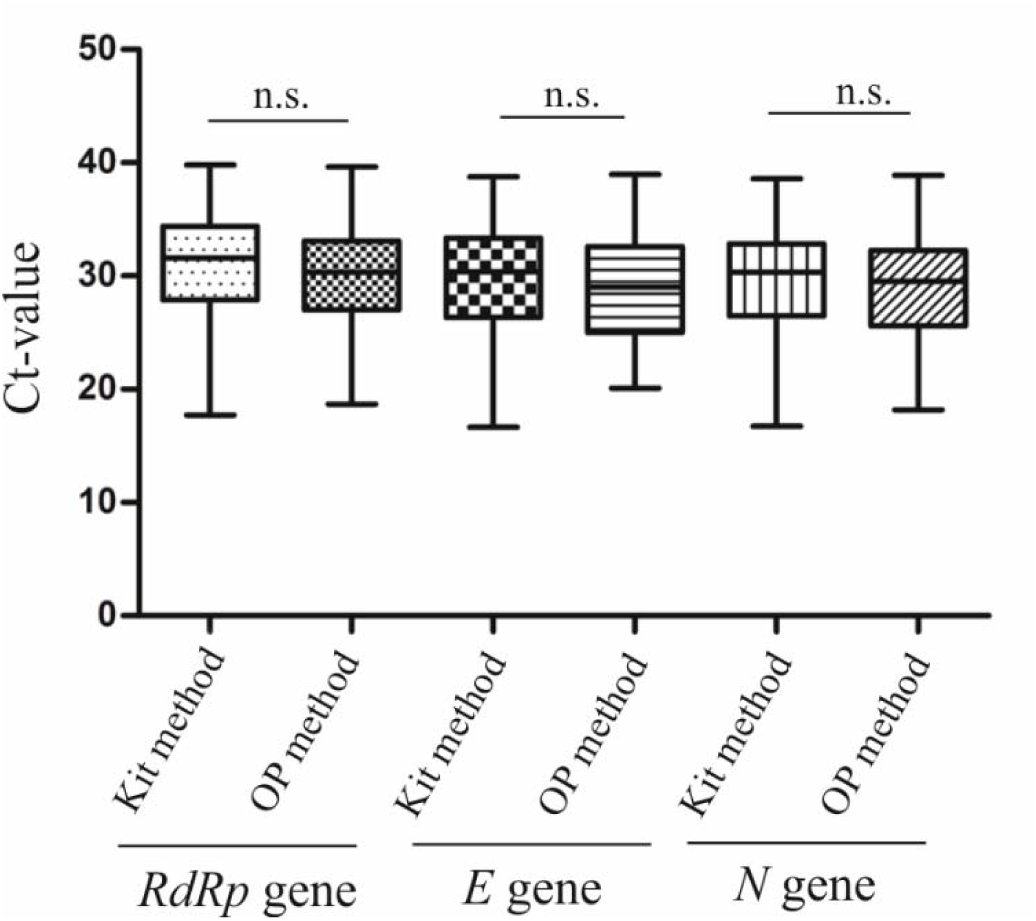
Box-plot showing no significant change in the Ct values of 250 COVID-19 infected patients (for *RdRp* gene, *E* gene and *N* gene) obtained from using the One-pot RNA isolation method when compared to Ct values acquired using commercially available RNA-isolation kits. (n.s.: not significant; Mann-Whitney test).

## Discussion

Globally, as on, 11 January 2022, there have been 308,458,509 confirmed cases of COVID-19, counting 5,492,595 deaths, reported to WHO (World Health Organization) (World Health Organization, 2022). However, in India, from 3rd January 2020 to 11 January 2022, there have been 35,875,790 confirmed cases of COVID-19 with 484,213 deaths, according to WHO (World Health Organization, 2022). From this 6 months study the prevalence rate of Covid-19 infection was found to be 18.52% in 2020 in Faridabad, Haryana India. Like previous data showing men are more susceptible to infection than women, our data also revealed that males are more predisposed to COVID-19 infection than females (Bwire, 2020). However, it was also reported by Jian-Min Jin et al. that men and women have the same Covid-19 prevalence rate but in the case of men, the risk of complications and death is much more, independent of age (Jin *et al*., 2020). Although previous studies pointed to the fact that COVID-19 prevalence increases in adolescents and youth compared to older adults, our data revealed that the rate of infection increased with the increasing age group of the individual suggesting that older people are more prone to COVID-19 infection than the younger ones (Rumain *et al*., 2021). The reason for this observation can be attributed to age-related gradual weakening of immune system function (immunosenescence) or maybe due to hyperactive yet ineffective immune system (inflammaging) in older people leading to systemic inflammation (FRANCESCHI *et al*., 2006; Aw *et al*., 2007).

To address various issues, several POCs have been established to enable the COVID-19 diagnosis outside the centralized testing facilities to hasten the clinical decision-making within a short window of time. Like, CRISPR-Cas9 enzymology-based diagnostic platform-specific high-sensitivity enzymatic reporter unlocking (SHERLOCK) provides ultra-high specific amplification of nucleic acid sequences from clinical samples with the use of an isothermal cycler (Kellner *et al*., 2019). Also, another CRISPR-based, FnCas9, Editor Linked Uniform Detection Assay (FELUDA) has been designed in India for rapid and accurate detection of SARS-CoV-2 nucleic acids at a very low cost (Azhar *et al*., 2021). In addition, a sensitive RNA-isolation protocol exploiting Trizol to purify viral RNA from various types of clinical specimens with nominal BSL-1 precautions has been formulated recently (Paz *et al*., 2020). However, this RNA-isolation method included many transfer steps and was time-consuming. To facilitate rapid, inexpensive and efficient testing of COVID-19 infected samples, we have tried to develop one tube RNA-isolation method which is capable of detecting SARS-CoV-2 RNA in the NP/OP swabs. The efficacy of this new method was tested by comparing the Ct values of the clinical samples with the Ct values obtained by employing commercially available RNA-isolation kits. The use of this new method is highly recommended when dealing with a huge number of clinical samples in low-resource settings like small mohalla clinics, primary health care centers, etc. This One-pot (OP) RNA-isolation method is highly cost-effective with a cost of approximately Rs. 35/sample when compared to commercially available RNA isolation kits with a cost of approximately Rs 200/sample. For isolating RNA, the time consumed by other commercial kits with the column method is more than 1 hour, but with this new method, the time required is approximately 30 mins. Thus, the use of this One-pot RNA isolation kit would highly benefit mankind by saving cost, time and labor thereby facilitating rapid and early detection of the disease. This study also implies the application of a resource-conserving testing algorithm employing sample pooling for clinical health diagnosis.

## Conclusion

This study from India characterizes the disease pattern and the possible outcomes associated with it. In this study, we have tried to investigate the spread of COVID-19 infection over the region of Faridabad, Haryana (northern India) over 6 months (May 2020-October 2020). The rate of infection was found to be higher in the case of individuals belonging to higher age groups. Also, we have successfully developed a crude RNA isolation method using a single tube which can be employed for testing a large number of samples in a low resource setting, without compromising the qRT-PCR results. This quick, sensitive and inexpensive RNA-isolation method can be a better-suited alternative to most of the expensive, time-consuming commercially available RNA isolation kits.

## Data Availability

All relevant data are within the paper. If required, the master chart of patients can be made available at reasonable request from the corresponding authors.

## Abbreviations

SARS-CoV-2: Severe Acute Respiratory Syndrome-Coronavirus-2
CoVs: Coronaviruses
COVID-19: Corona Virus Disease-19
MERS: Middle East Respiratory Syndrome
VTM: Viral Transport Medium
POC: Point of Care
OP method: One-Pot method
DEPC: Diethyl Pyrocarbonate
NP/OP: Nasopharyngeal/Oropharyngeal
WHO: World Health Organization
CRISPR: Clustered Regularly Interspaced Short Palindromic Repeats
ICMR: Indian Council of Medical Research
SARI: Severe Acute Respiratory Illness
ILI: Influenza Like Illness
Ct-value: Cycle threshold value
BSL-1: Biosafety Level-1
USA-CDC: United States of America-Centers for Disease Control and Prevention
qRT-PCR: quantitative Reverse Transcription-Polymerase Chain Reaction
*N* gene: Nucleocapsid gene
*RdRp* gene: RNA dependent RNA polymerase gene
*E* gene: viral Envelope gene

## Authors’ contributions

Conceptualization, Sonia Jain, Arghya Bhowmick and Anil Pandey; Formal analysis, Sonia Jain and Arghya Bhowmick; Funding acquisition, Anil Pandey; Investigation, Sonia Jain and Arghya Bhowmick; Methodology, Sonia Jain, Arghya Bhowmick and Anil Pandey; Project administration, Sonia Jain and Anil Pandey; Supervision, Sonia Jain and Anil Pandey; Writing – original draft, Sonia Jain and Arghya Bhowmick; Writing – review & editing, Sonia Jain, Arghya Bhowmick and Anil Pandey.

## Acknowledgements

The authors would like to thank ESIC Medical College & Hospital, Faridabad, Haryana, India for providing intramural funding to SJ. AB is supported by a fellowship from CSIR (Council of Scientific and Industrial Research), Government of India [File No. 09/015(0525)/2017-EMR-I].

## Ethics approval and consent to participate

Ethical clearance was obtained from the Ethical committee of ESIC medical college and hospital, Faridabad before conducting the study, and informed oral consent from the patients was taken. All the necessary protocols required for COVID-19 diagnosis were followed according to ICMR (Indian Council of Medical Research) guidelines.

## Consent for publication

Not applicable

